# Modeling the COVID-19 pandemic - parameter identification and reliability of predictions

**DOI:** 10.1101/2020.04.07.20056937

**Authors:** Klaus Hackl

## Abstract

In this paper, we try to identify the parameters for two elementary epidemic models, the so-called SI- and SIS-models, via non-linear regression using data of the COVID-19 pandemic. This is done based on the data for the number of daily infections. Studying the history of predictions made, we attempt to estimate their reliability concerning the future course of the epidemic. We validate this procedure using data for the case numbers in China and South Korea. Then we apply it in order to find predictions for Germany, Italy and the United States. The results are encouraging, but no final judgment on the validity of the procedure can yet be made.

## 1 Introduction

In most countries, social distancing measures are in effect now in order to fight the COVID-19 pandemic. Considering the serious effects of these measures on the affected societies and the ensuing political discussions on their intensity and duration, it would be highly desirable to be able to make modeling based predictions on the future timeline of the epidemic, as long as the measures are upheld. Of course, many attempts are made in this direction. However, most of them require very detailed data that are laborious and time-consuming to generate.

In this work, we try to study the possibility to base predictions on data sets readily available, namely the number of reported infections. We are aware, that these numbers depend strongly on the intensity of testing done in the various countries and the reliability of the reported numbers. In this work we presume that there is a factor, country-specific, but constant in time, between the reported and the actual number of cases. If this assumption were valid, the total number of infected individuals would be off by this very factor. However, other parameters, like the point in time when the peak in the numbers of daily infections would occur, or the following rate of decay of these numbers, would not be affected.

This is an updated version of the preprint [2]. The following changes have been made:

We use now the the number of daily infections averaged over seven days. This eliminates the observed effects of periodic delays in reporting of cases and leads to much more regular data, thus that local minimization can be used for parameter identification. We abandoned the use of accumulated cases as done in [2], since these data proved very insensitive to variation of the model parameters.

It turned out, that often the decrease in the number of daily infections after the peak is slower than the increase observed before. This effect can be captured nicely by extending the SI-model employed in [2] to the so-called SIS-model. This also allows a preliminary judgment of the effectiveness of the social distancing measures taken.

Since a prominent decrease in the number of daily infections can be observed in some countries now, we can use these data to perform a post-analysis of the reliability of the predictions made based on the procedure presented in this paper.

Finally, we would like to stress, that we intend this work to be the starting point of a discussion and maybe further research. By no means, having a background in engineering and not in virology or epidemiology, we are claiming any medical expertise. The paper should be rather seen as a general exercise in modeling and interpretation of data.

## 2 An elementary model

Our aim is to model a situation where social distancing measures are in effect, as currently is the case in most countries. This means, that only a small portion of the population is affected, which is well but not completely isolated from the rest. As starting point, we refer to the SIS-compartmental model, see [5], compare also the SIR-compartmental model, [4]. It is defined by the differential equations

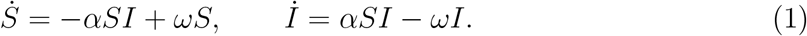

Here *I*(*t*) is the number of individuals in the infectious population and *S*(*t*) denotes the number of individuals in the susceptible population, in our case those who can get infected because they are not protected by social distancing. The parameter *α* is related to the effective reproduction number by

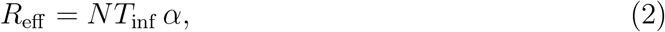

where *N* is the initially susceptible population and *T*_inf_ is the time period during which an individual is infectious. For Sars-Cov-2, no definite value for *T*_inf_ has yet been reported. The parameter *ω* defined in our case the rate of exchange of individuals between the contained part of the population and the rest. Hence, it can be considered a measure of the effectiveness of the social distancing.

The system of ordinary differential equations given by Eqs. (1) possesses the closed form solution

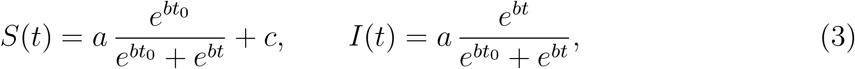

Where

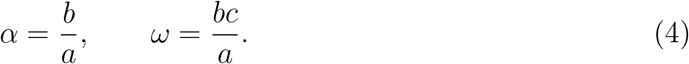

Note that parameter *a* does not correspond to *a* in [2]. Obviously, we have

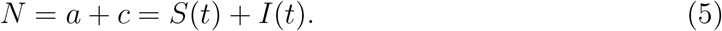

The instant given by *t*_0_ marks in some sense the peak of the epidemic, defined by

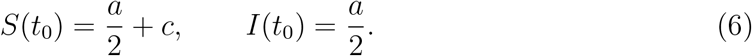

We have the initial conditions

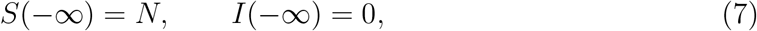

and the limiting values

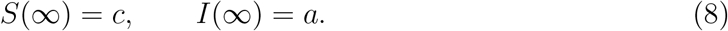

For the purpose of parameter identification, we need the cumulative number of infected individuals *I*_c_, defined as

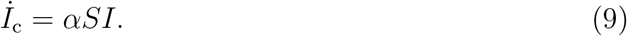

Substitution of Eqs. (3) and (4) into Eq. (9) gives

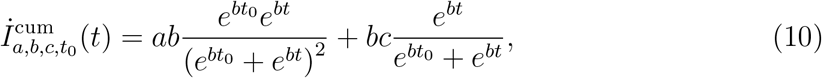

and

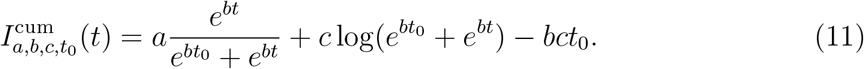

## 3 Parameter identification

We determine the three parameters of our model via non-linear regression. The data taken from the worldometer web page, [1], which essentially uses the data from the Johns Hopkins University Center for Systems Science and Engineering (JHU CCSE). For the parameter identification done in this paper, we have used the available data up to including Apr. 20, 2020. The data are provided in form of lists 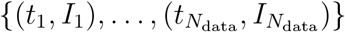 for the total number of infections up to day *t*_*i*_, and 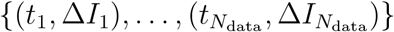 for the number of daily infections. Time is measured in days, starting on Jan. 1, 2020. Hence, *t* = 1 d corresponds to Jan. 1, *t* = 32 d to Feb. 1, *t* = 61 d to Mar. 1, 2020, and so on. Obviously, we have

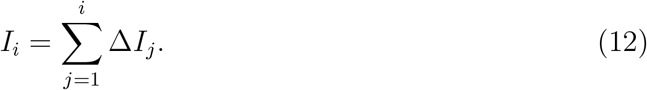

In [2] we employed the number of total infections as well for parameter identification. However, we noticed that this leads to rather ill-conditioned problems giving results with little predictive power. So we did not pursue this approach anymore.

In order to achieve stable parameter identification, we use 7-day averages of the data, eliminating periodic oscillations caused by delays in data reporting. The *n*-day average is given as

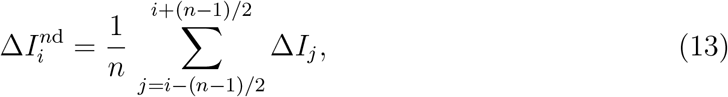

for *n* odd. As an example, we show the result of this procedure for the data concerning South Korea and Germany in Fig. 1. Note that the lists of averaged data are shorter by three days at their beginning and end.

**Figure 1:**
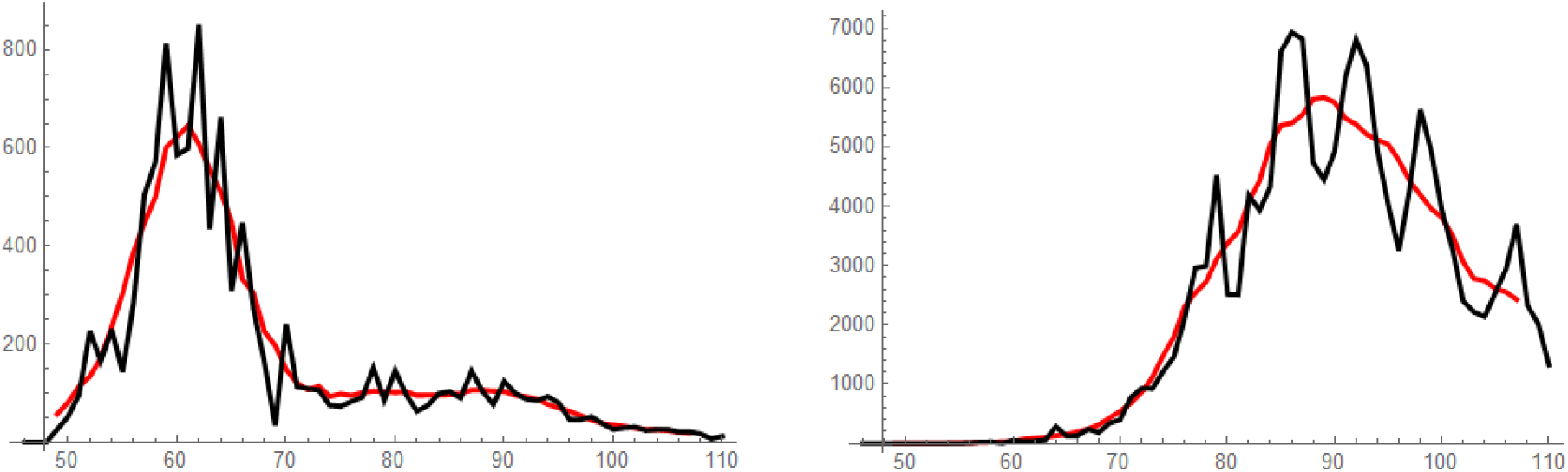
Daily cases Δ*I*_*i*_ in black and 7-day averages 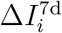 in red, South Korea left,Germany right.

The model function corresponding to the averaged data over an interval Δ*t* is given by

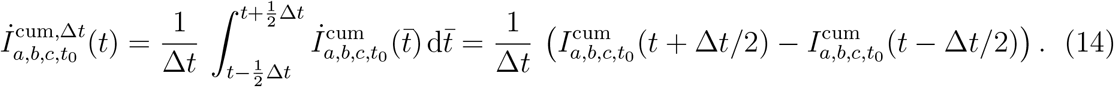

In order to identify the model parameters, let us define the error *e*_7d_(*a, b, c, t*_0_) with respect to 7-day averages of daily cases by

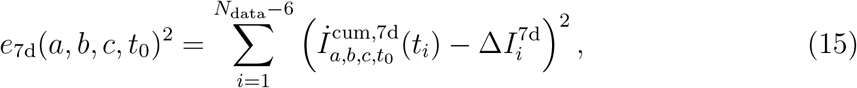

the data norm

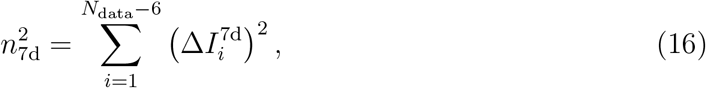

and the relative error

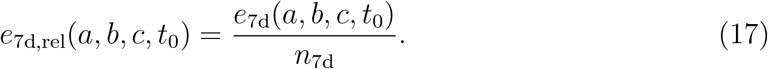

Finally, we find the parameters *a, b, c, t*_0_ by minimizing the errors. However, it turns out, that the determination of parameter *c* is rather ill-posed during stages of the epidemic, when the number of daily infections is still on the rise. This effect often leads to overly pessimistic predictions. Our solution to this problem is to reduce the SIS-model to the so-called SI-model during this stage, setting *c* = 0. Thus, we are performing two minimization procedures:

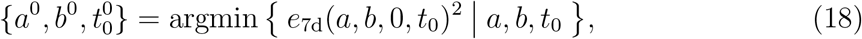

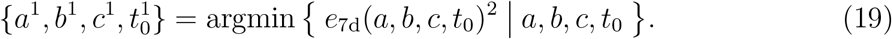

Minimization is done using the computer algebra system *Mathematica*, [3]. For our purposes, the local minimization algorithm given by the *FindMinimum* function, which uses a version of an Newton-Raphson procedure, works fine. Attention has to be given, though, to choosing appropriate initial values for the parameters in order to achieve convergence.

Both parameter identification procedures given in Eqs. (18) and (19) are performed every time. Then the parameter set {*a*^1^,*b*^1^,*c*^1^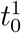} will be preferred over {*a*^1^,*b*^1^,*c*^1^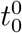} as soon as it returns a significantly smaller error defined as

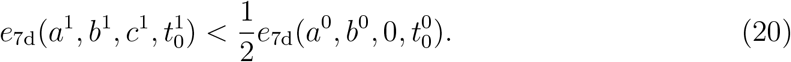

## 4 Results

In Figs. 2 to 6, the 7-day averages of the numbers of daily cases (left) and the non-averaged numbers of total cases (right) are plotted versus time in days. The corresponding data are shown in blue color.

**Figure 2:**
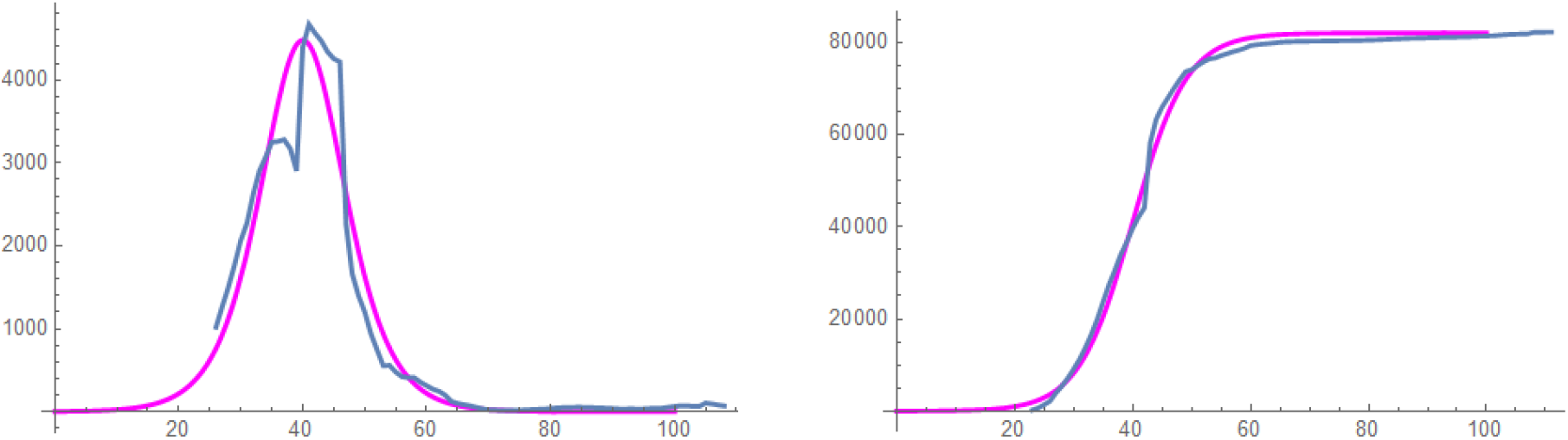
China, left: 7-day average of daily cases, 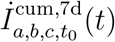 (magenta) versus data 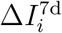 (blue), right: total cases 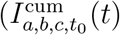 (magenta) versus data *I*_*i*_ (blue).

In Fig. 2 and Fig. 3 the data for China and South Korea are displayed. Both countries can be considered to be in a late stage of the epidemic and the data are matched well by the model. In Figs. 4, 5 and 6, the corresponding graphs are plotted for Germany, Italy and the United States. These countries can be considered to be in earlier stages of the epidemic. In these cases, the modeling agrees very well with the data, too. Interestingly, a prominent decline in daily cases is predicted for Germany, while for Italy and the United States, the daily cases are predicted to remain at a high level. However, we would rather argue, that this means, that a reliable prediction is not possible from the data at this stage.

**Figure 3:**
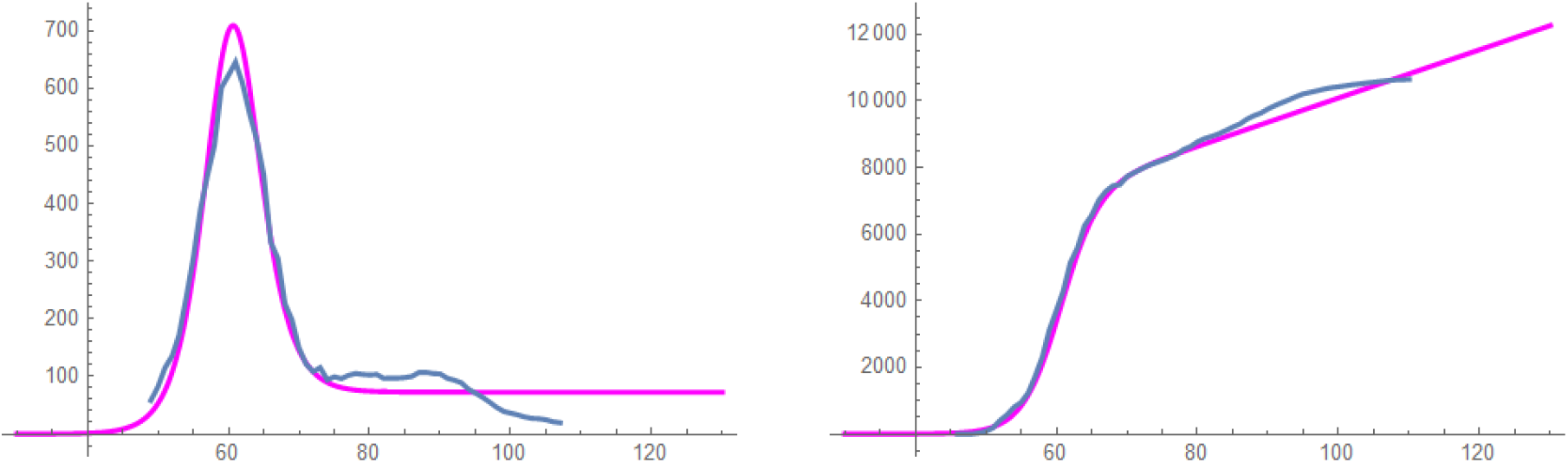
South Korea, left: 7-day average of daily cases, 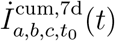 (magenta) versus data 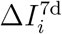 (blue), right: total cases 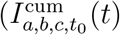 (magenta) versus data *I*_*i*_ (blue).

**Figure 4:**
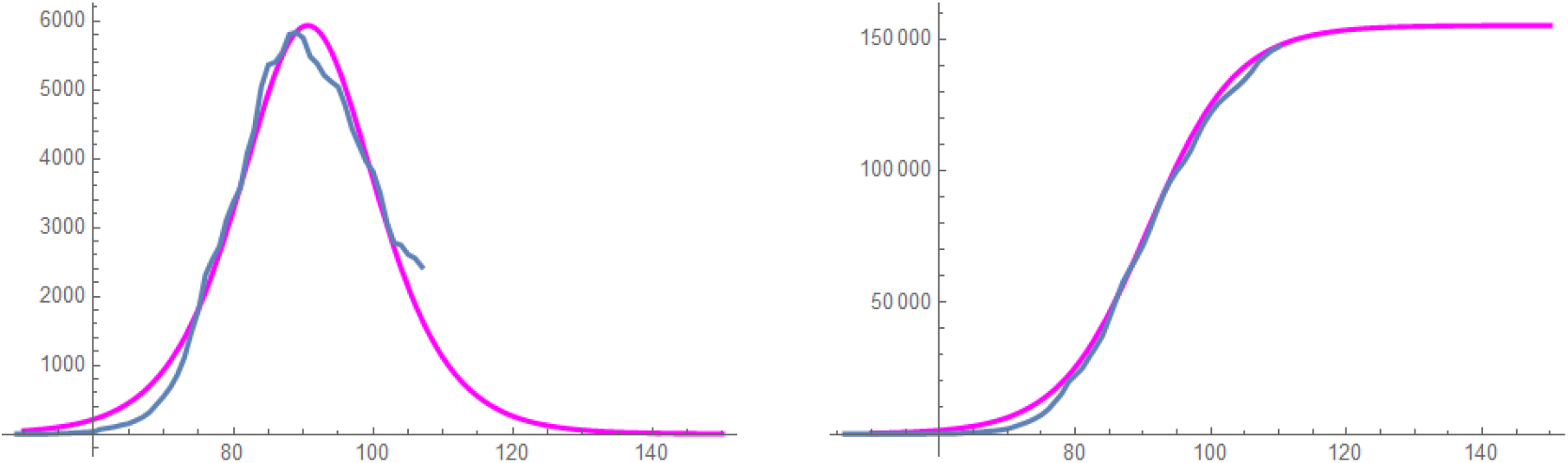
Germany, left: 7-day average of daily cases, 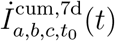 (magenta) versus data 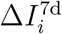 (blue), right: total cases 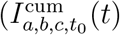 (magenta) versus data *I*_*i*_ (blue).

Some key data provided by the model are given in Table 1.

**Table 1:** values of key parameters on Apr. 20, 2020

| | $a$ | $b$ | $c$ | $t_0$ | $e_{7d,rel}$ |
| --- | --- | --- | --- | --- | --- |
| China | 81970 | 0.22 | 0 | 40.0 | 0.20 |
| South Korea | 7225 | 0.37 | 195 | 60.5 | 0.09 |
| Germany | 155134 | 0.15 | 0 | 90.7 | 0.09 |
| Italy | 82430 | 0.19 | 17956 | 90.7 | 0.03 |
| United States | 251413 | 0.22 | 129713 | 91.1 | 0.02 |

## 5 Reliability of predictions

Let us attempt a post-analysis here. In Figs. 7, 9, 11, 13, and 15, we show the development of the identified model parameters *a, b, c, t*_0_ over time. And in Figs. 8, 10, 12, 14, and 16, the model results obtained at several instances in time a displayed versus the 7-day averaged numbers of daily infections. Black dots indicate the specific instances in time. Hence, the available data have been employed up to this point.

**Figure 5:**
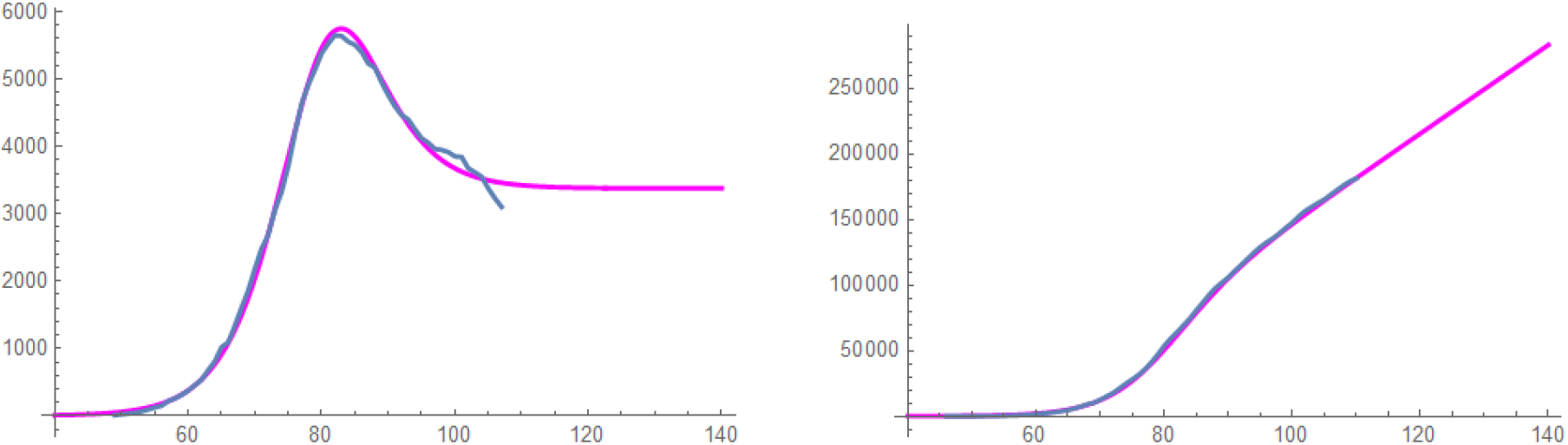
Italy, left: 7-day average of daily cases, 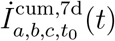 (magenta) versus data 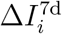 (blue), right: total cases 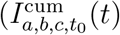 (magenta) versus data *I*_*i*_ (blue).

**Figure 6:**
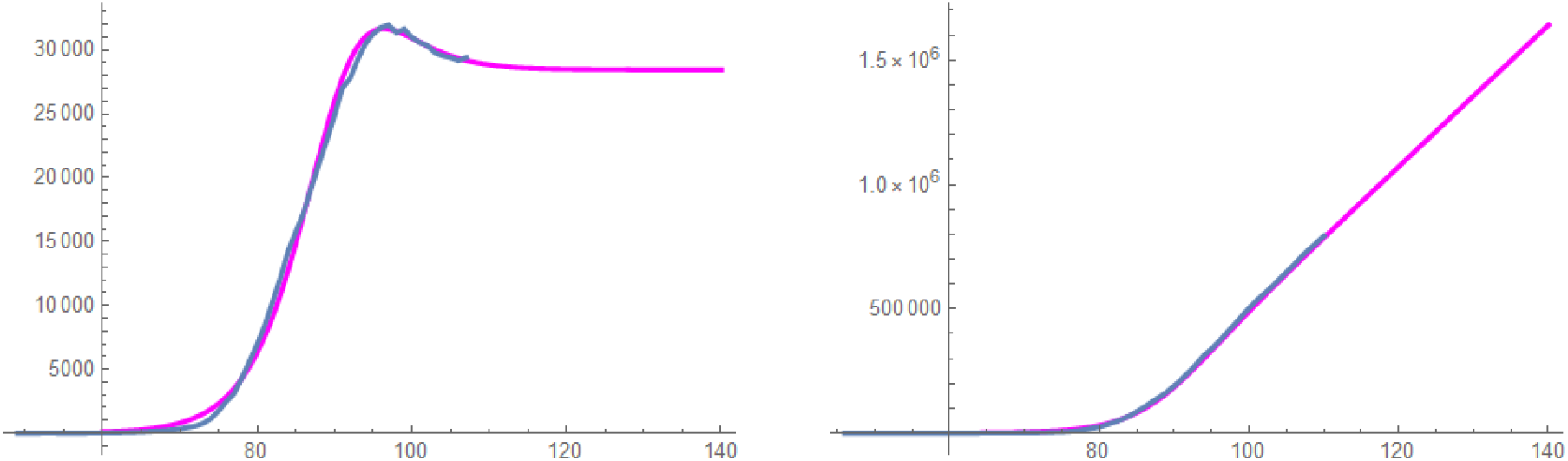
United States, left: 7-day average of daily cases,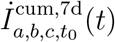 (magenta) versus data 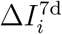 (blue), right: total cases 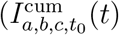 (magenta) versus data *I*_*i*_ (blue).

**Figure 7:**
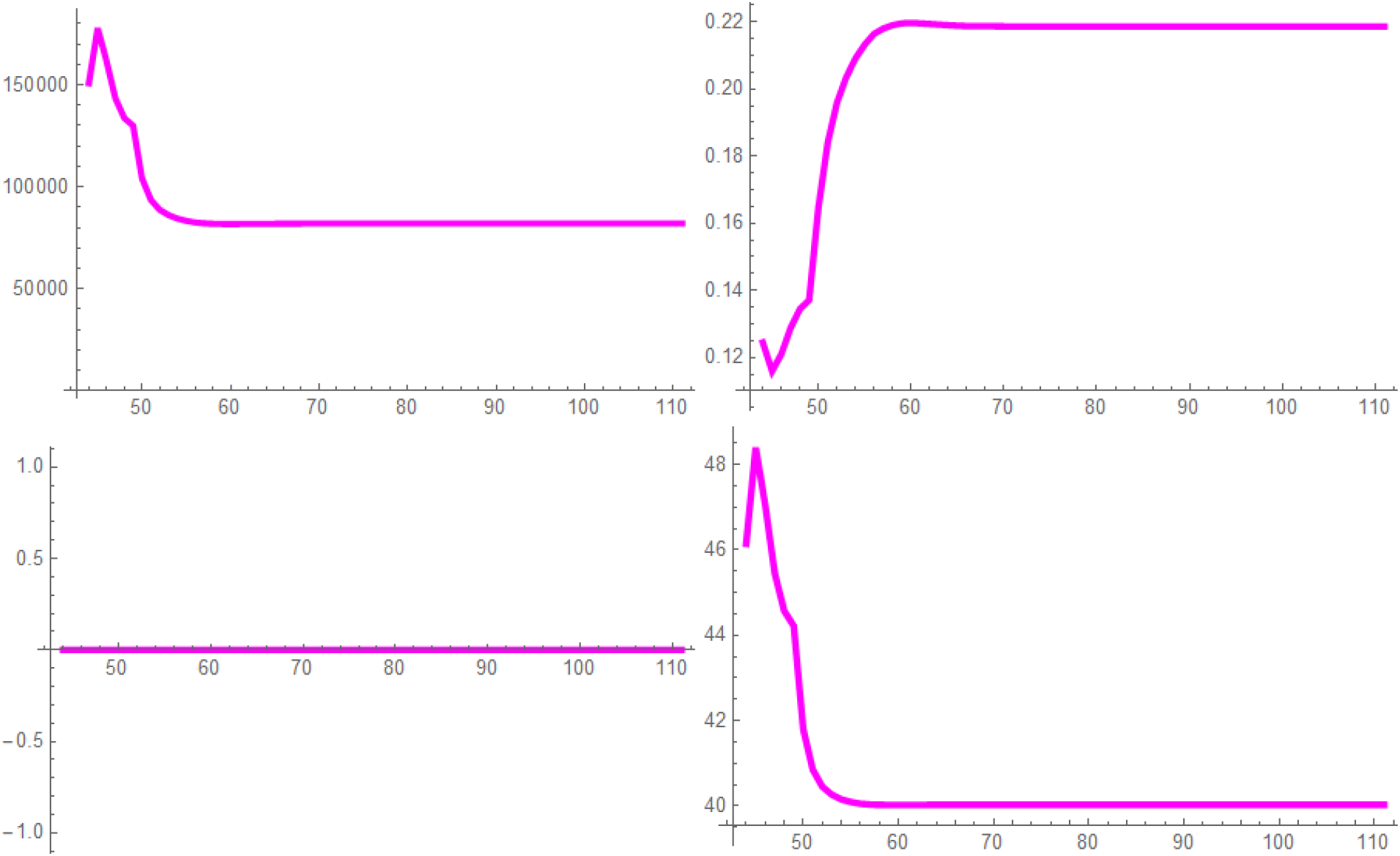
China, model parameters *a* (upper left), *b* (upper right), *c* (lower left), *t*_0_ (lower right) versus time in days.

**Figure 8:**
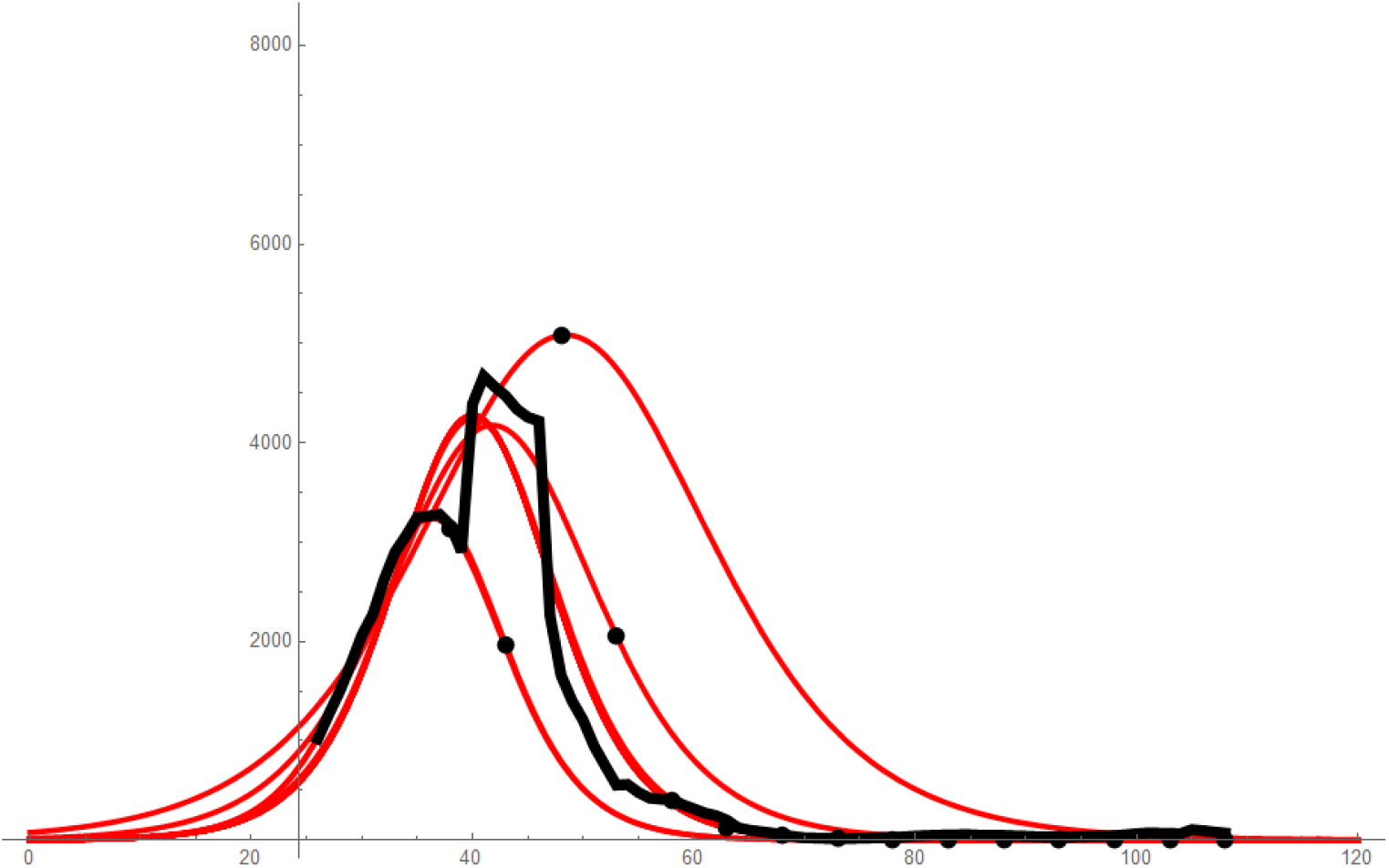
China, timeline of predictions of daily infections, 7-day averaged data in black,model predictions in red, black dots mark the point in time up to which data have been used.

**Figure 9:**
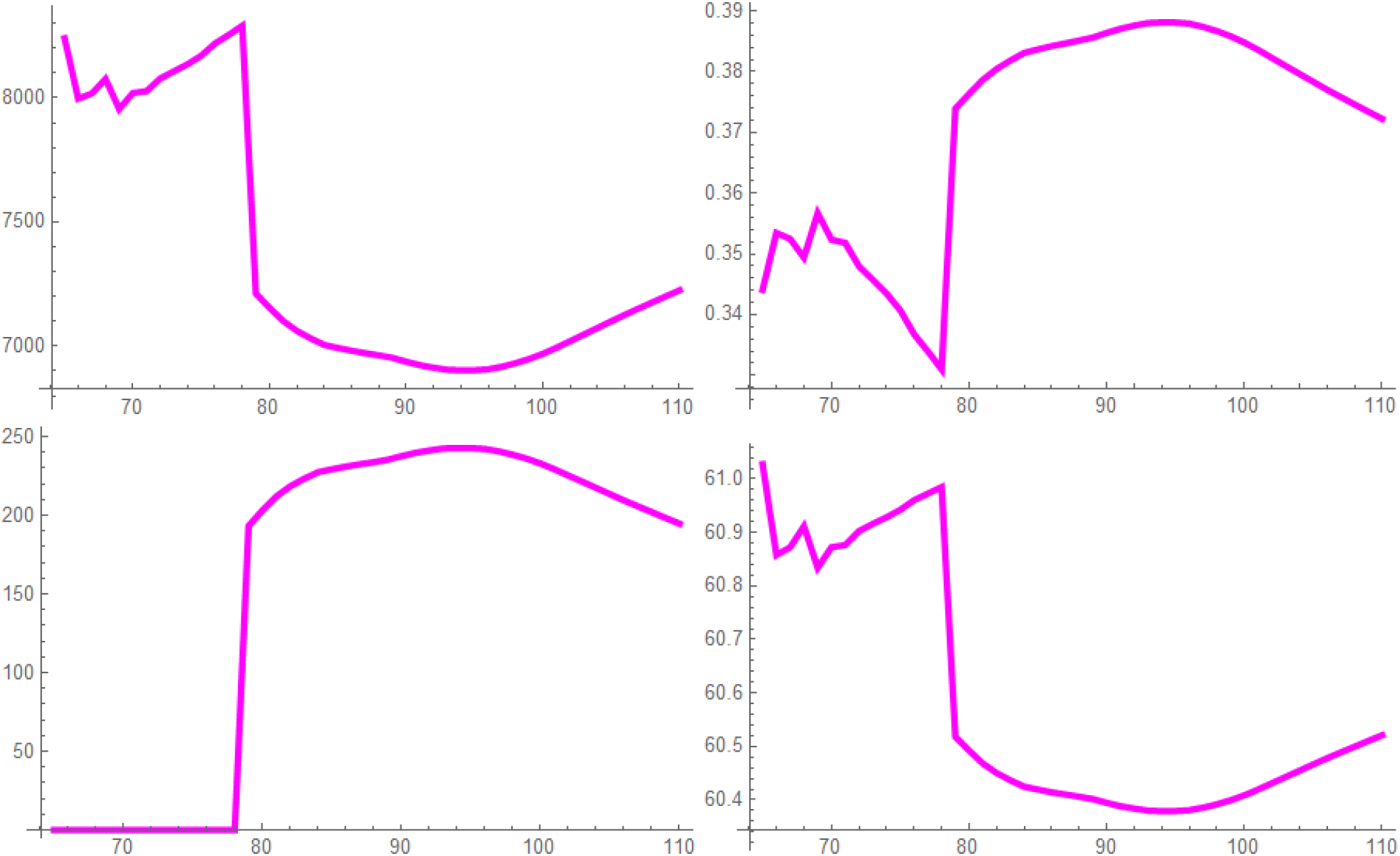
South Korea, model parameters *a* (upper left), *b* (upper right), *c* (lower left), *t*_0_ (lower right) versus time in days.

**Figure 10:**
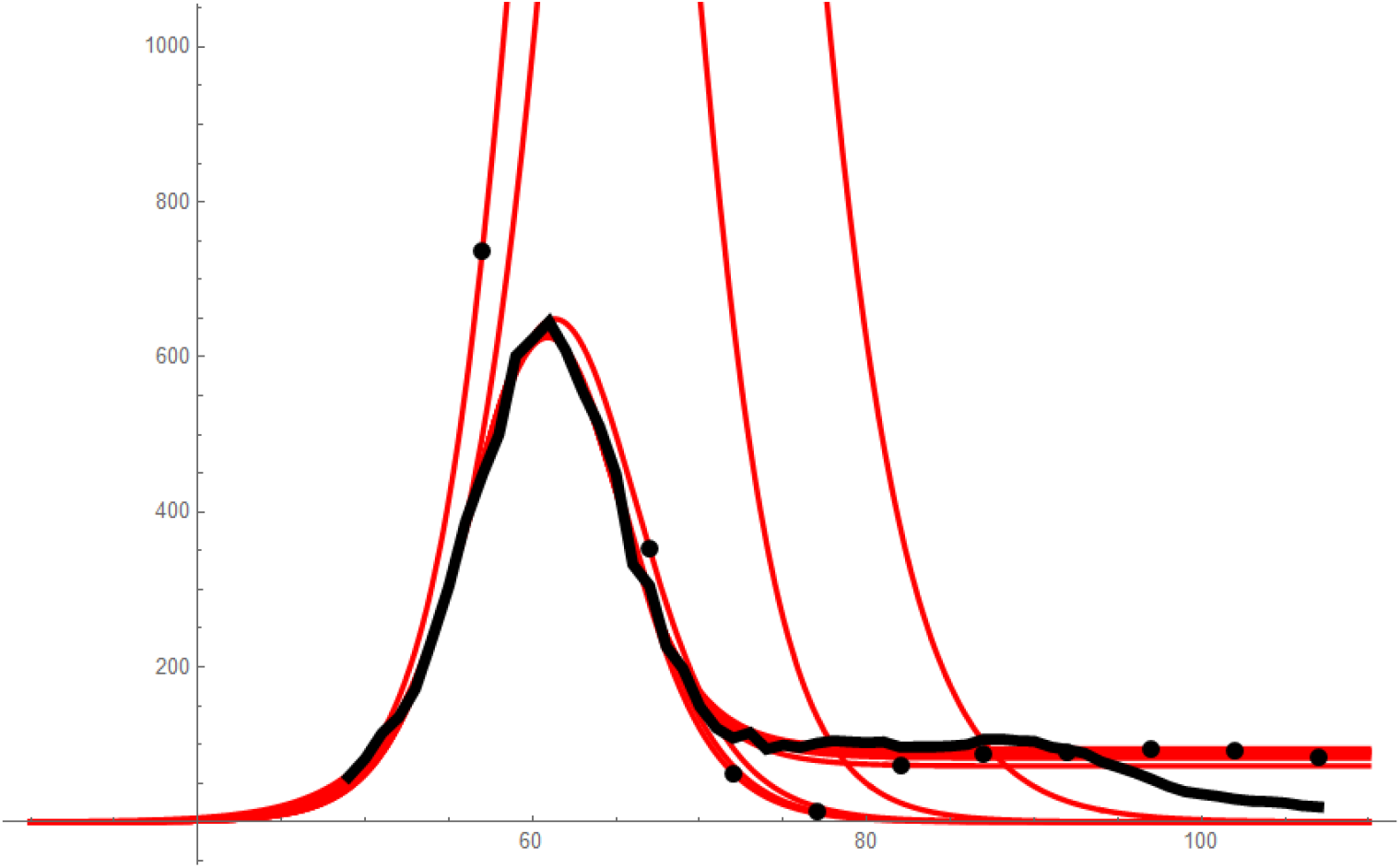
South Korea, timeline of predictions of daily infections, 7-day averaged data in black,model predictions in red, black dots mark the point in time up to which data have been used.

**Figure 11:**
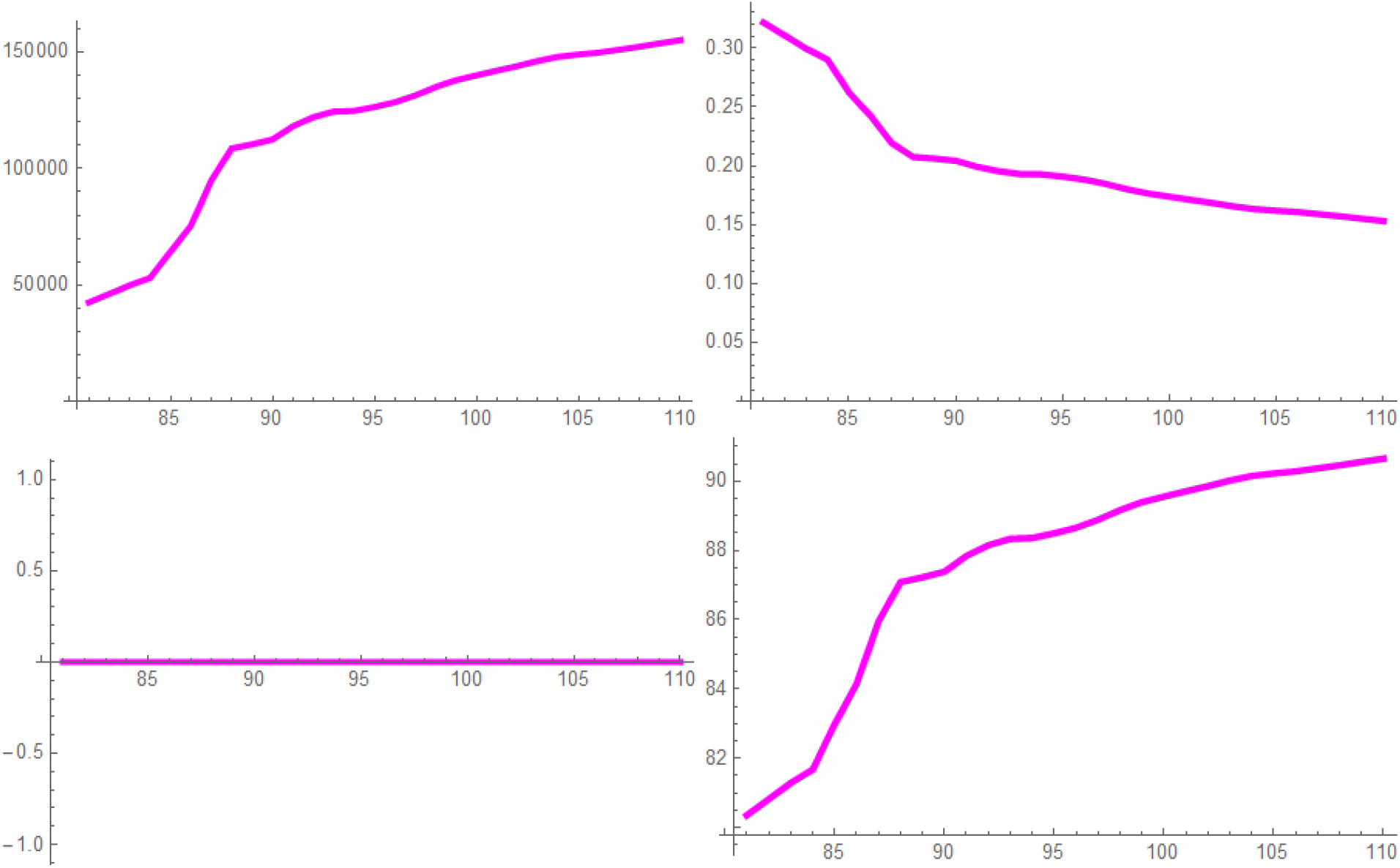
Germany, model parameters *a* (upper left), *b* (upper right), *c* (lower left), *t*_0_ (lower right) versus time in days.

**Figure 12:**
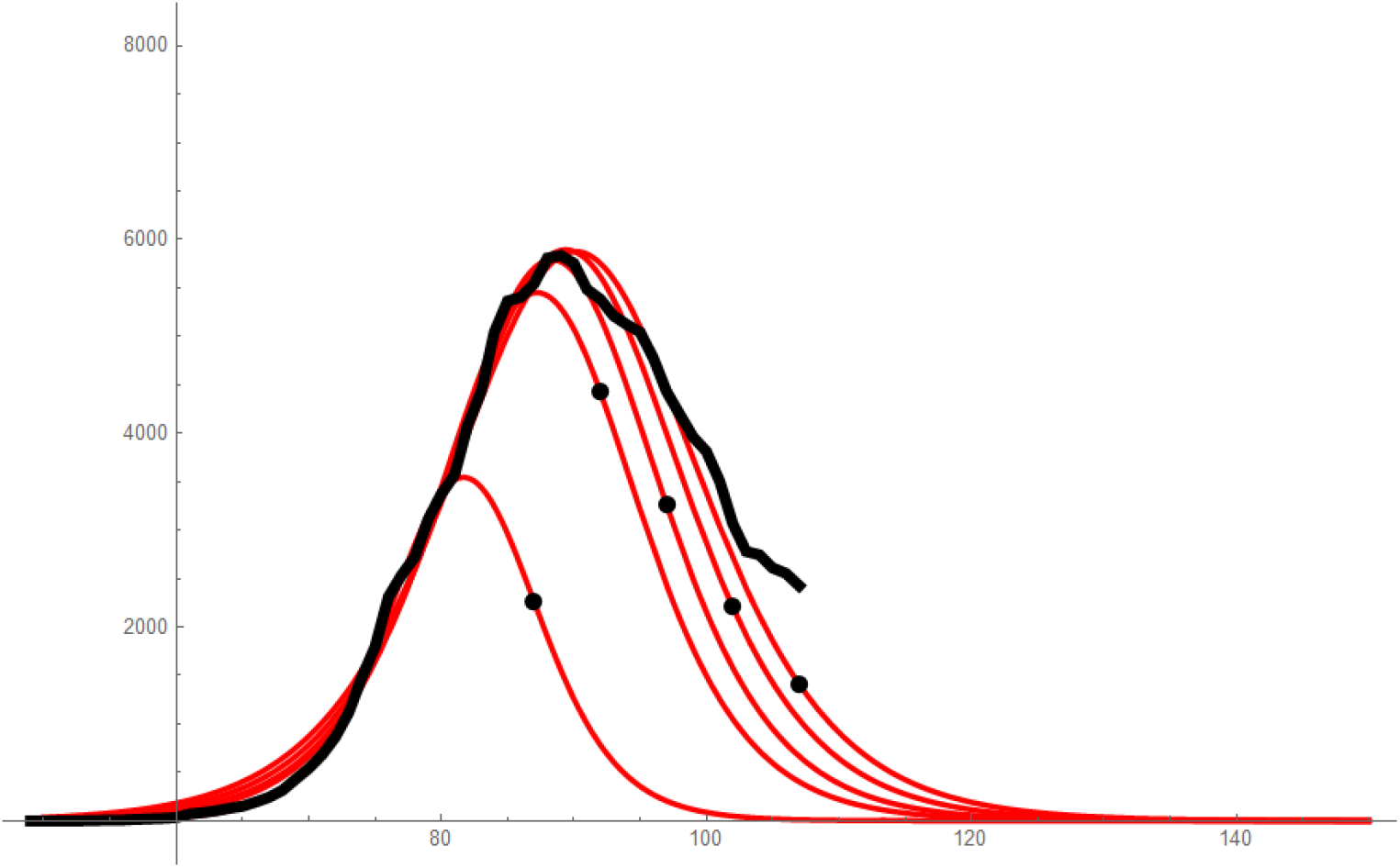
Germany, timeline of predictions of daily infections, 7-day averaged data in black,model predictions in red, black dots mark the point in time up to which data have been used.

**Figure 13:**
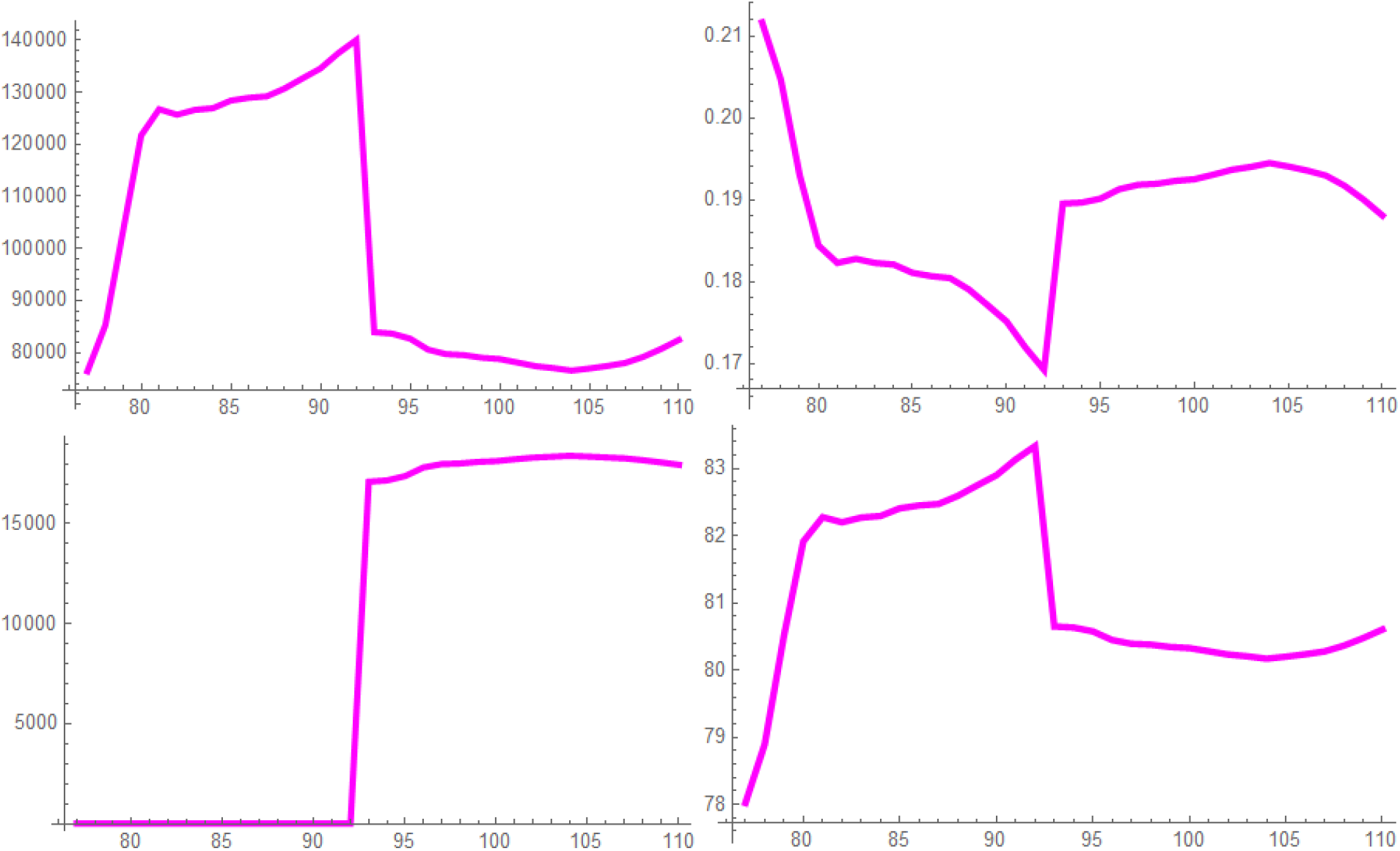
Italy, model parameters *a* (upper left), *b* (upper right), *c* (lower left), *t*_0_ (lower right) versus time in days.

**Figure 14:**
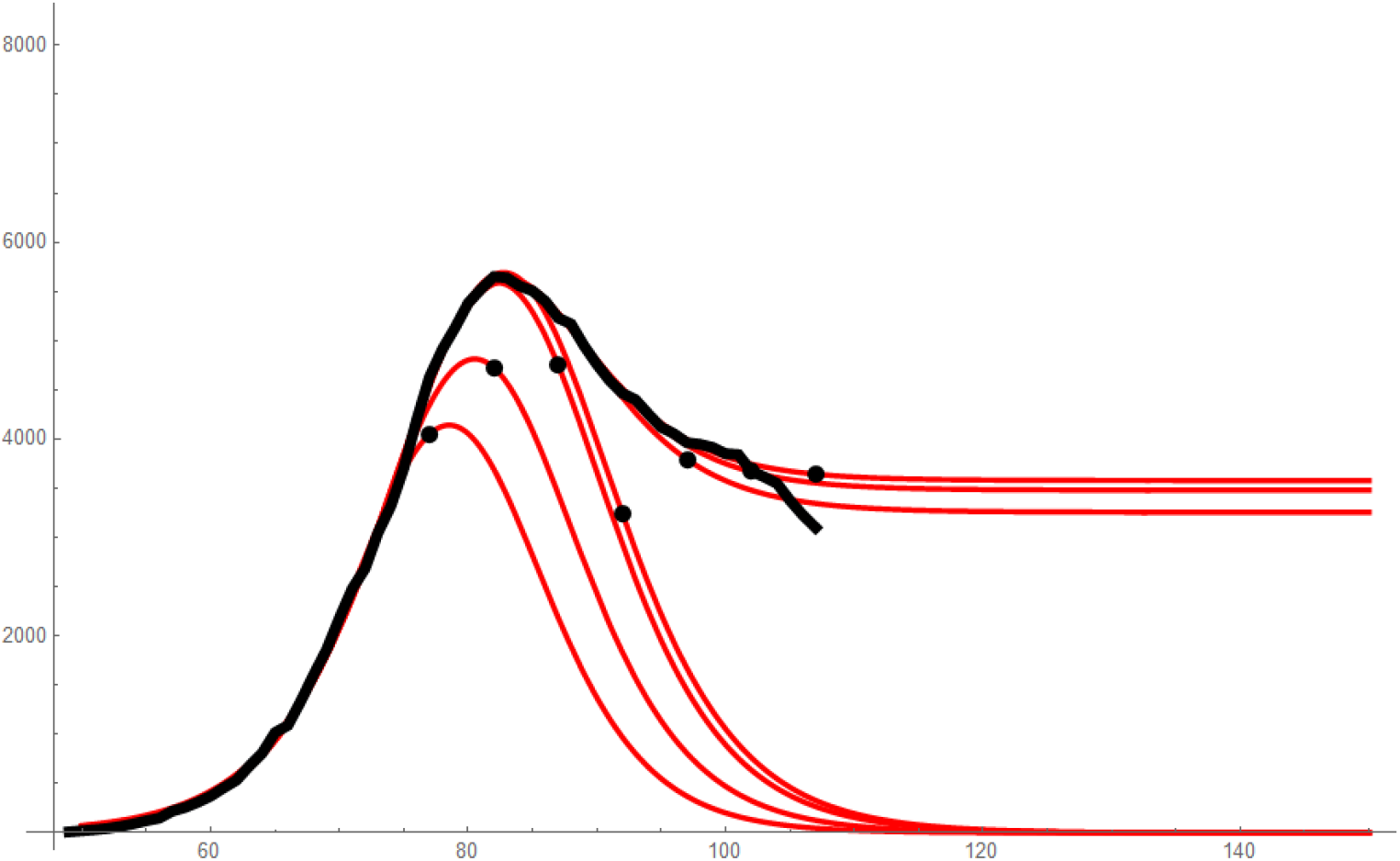
Italy, timeline of predictions of daily infections, 7-day averaged data in black,model predictions in red, black dots mark mark the point in time up to which data have been used.

**Figure 15:**
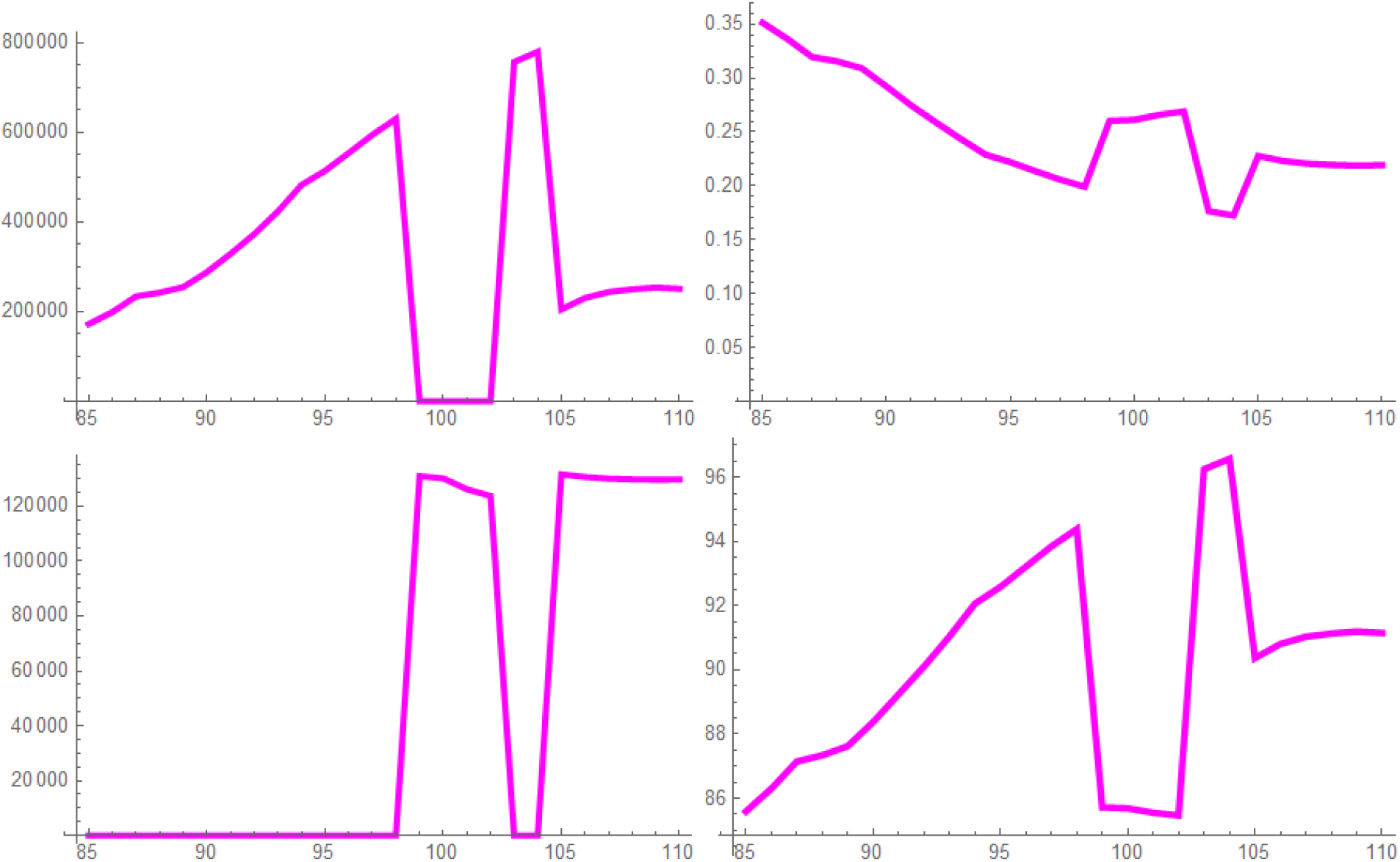
United States, model parameters *a* (upper left), *b* (upper right), *c* (lower left), *t*_0_ (lower right) versus time in days.

**Figure 16:**
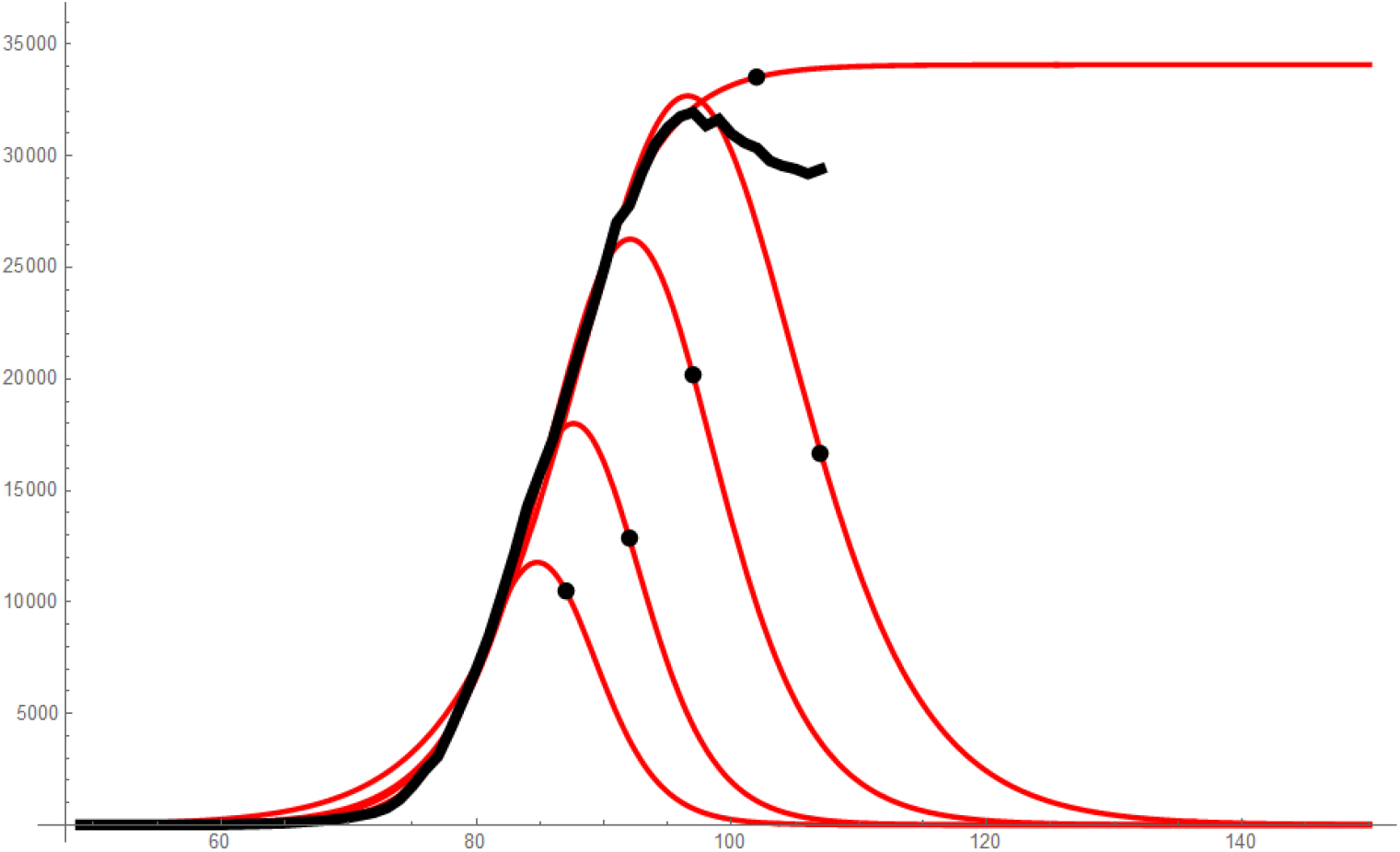
United States, timeline of predictions of daily infections, 7-day averaged data in black,model predictions in red, black dots mark the point in time up to which data have been used.

For China (Figs. 7, 8), it can be seen that the model parameters remain constant after *t* = 56d. This coincides with the point where the model starts to agree with the data. Before this instance, the number of infections is either under- or overestimated. Unfortunately, we have to state low predictivity here.

For South Korea (Figs. 9, 10), the model parameters vary only a little after *t* = 78d. Once again, this coincides with the point where the model starts to agree with the data. However, in comparison to China, this instant is reached earlier in time, though in the declining phase of the epidemic.

For Germany (Figs. 11, 12), we have converged parameters past *t* = 95d. We can see, that we have reliable predictions shortly after the peak in the number of daily infections. The predictions are on the optimistic side with respect to the total duration of the epidemic.

An interesting development of the predictions can be observed for Italy (Figs. 13, 14). At first, the parameters behave similarly as in the case of Germany and the predictions appear to converge. Then, however, the parameter *c* jumps suddenly to a finite value, because the number of daily infections declines slower than expected. At the moment, it can not be said at which level this number will stabilize.

For the United States (Figs. 15, 16), the parameter *c* fluctuates between zero and a finite value, indicating a very dynamic course of the epidemic. No reliable predictions can be made yet.

## 6 Conclusion

We have identified the parameters in an elementary epidemic model via non-linear re-gression using data of the covid-19 pandemic. Furthermore, we have attempted to get an insight into the reliability of predictions based on this procedure by observing the time-line of the parameters calculated. Our results indicate, that this approach might work. However, reliable predictions seem to be possible after the peak of the number of daily infections has already been reached. For earlier predictions, a better understanding of the internal mechanisms of the epidemic is probably required. This has to go along with the availability of more detailed data than just the plain numbers of daily infections.

## Data Availability

Data for the COVID-19 pandemic available at the following web sites have been used.

https://www.worldometers.info/coronavirus/

https://data.humdata.org/dataset/novel-coronavirus-2019-ncov-cases

